# Surgical interventions targeting the nucleus caudalis for craniofacial pain: a systematic and historical review

**DOI:** 10.1101/2022.01.04.22268756

**Authors:** Brendan Santyr, Mohamad Abbass, Alan Chalil, Amirti Vivekanandan, Margaret Tindale, Nicholas M. Boulis, Jonathan C. Lau

**Author notes:** ***Corresponding Author:*** Jonathan C. Lau, M.D., Ph.D., FRCSC Department of Clinical Neurological Sciences University Hospital, 339 Windermere Rd, London, Ontario, Canada N6A 5A5. ***Disclosure of funding:*** No funding was received for the completion of this work.

## Abstract

**Introduction:** Craniofacial pain is a prevalent group of conditions and when refractory to conventional treatments poses a significant burden. The last decade has seen a renewed interest in the multimodal management of pain. Interventions targeting the nucleus caudalis (NC) of the trigeminocervical complex have been available as a treatment option since the 1930s, yet evidence for efficacy remains limited.

**Methods:** We present a systematic review of the literature providing a historical perspective on interventions targeting the NC leading up to the present. We examine the various intervention techniques, clinical indications, and procedural efficacy. A novel outcome reporting scheme was devised to enable comparison among studies due to historically variable reporting methods.

**Results:** A review of the literature revealed 33 retrospective studies published over the last 80 years reporting on 827 patients. The most common technique was the open NC dorsal root entry zone nucleotomy/tractotomy; however, there has been an emergence of novel approaches such as endoscopic and spinal cord stimulation in the last 10 years. Regardless of intervention technique or preoperative diagnosis, 87% of patients demonstrated improvement with treatment.

**Conclusion:** The literature surrounding NC intervention techniques is reviewed. Recent advancements and the wide range of craniofacial pain syndromes for which these interventions show potential efficacy is discussed. New and less invasive techniques continue to emerge as putative therapeutic options. However, prospective studies are lacking. Furthermore, the evidence supporting even well-established techniques remain of poor quality. Future work should be prospective, utilize standard outcome reporting, and address efficacy comparisons between intervention type and preoperative diagnosis.

## Introduction

Refractory craniofacial pain is a heterogeneous group of disorders with multiple classification systems depending on clinical features that include anatomical distribution, anatomical origin, pathology, and symptomatology.^1^ The prevalence of craniofacial pain is estimated to be 26%.^2^ Epidemiological studies have yet to define the social and economic burden of this complex and debilitating group of diseases, however estimates may be made from the literature on trigeminal neuralgia (TN), the best studied of the chronic craniofacial pain syndromes outside the realm of headache. TN has been reported to impact multiple health status domains including activity, mood, ambulation, work, relationships, sleep, and life enjoyment.^3^ More than one-third of patients with TN had employment impacted through work hour reduction, disability, or unemployment.^3^ Despite advancements over the past two decades, many TN patients remain refractory to medical and conventional surgical options.^4^ Conventional surgical approaches to treat TN have targeted the nerve root and ganglion with demonstrated efficacy in treating typical paroxysmal TN type 1 pain but are relatively ineffective at treating the background pain characteristic of TN type 2.^5^ Worsening of constant background pain and the development of neuropathic pain can be seen with repeated treatments targeting the anterior system (distal to the nerve root).^6^ Therefore, an investigation into alternative surgical approaches and treatment options for these syndromes remains crucial to improving pain management.

Nociceptive craniofacial sensation, temperature, and crude touch are carried by general somatic afferent first-order neurons of the Vth, VIIth, IXth, and Xth cranial nerves to the trigeminocervical nuclear complex, where they synapse to second-order neurons within the nucleus caudalis (NC).^7^ The trigeminal nucleus forms ascending connections to structures involved in processing nociceptive stimuli, such as the thalamus, hypothalamus, locus coeruleus, and periaqueductal gray. Here nociceptive stimuli are modulated and pain is perceived.^8^ There is partial functional overlap between the C2 and C3 substantia gelatinosa and the NC, and it is this close anatomical relationship that inspired the use of dorsal root entry zone (DREZ) lesioning techniques to treat craniofacial pain.^8^

Interruption of the trigeminocervical complex at the level of the brainstem was first reported in humans in 1938.^9^ Introduced in 1968, DREZ procedures for chronic pain have progressed to include various methods of lesioning or modulating the DREZ at the spinal level of the pain, attempting to eliminate hyperactivity and spontaneous discharges that result in central perception of pain.^10–13^ Favorable outcomes in DREZ operations for other pain syndromes and advancements in techniques for open and percutaneous trigeminal tractotomy helped provide motivation for NC DREZ procedures for craniofacial pain.^14–17^ Similar to standard spinal DREZ procedures that target the dorsal horn of the spine, NC DREZ operations target the spinal trigeminal nucleus pars caudalis at the cervicomedullary junction, relieving pain through destruction of the second-order neurons.^10,17–19^

Although the first application of lesioning of the trigeminocervical complex dates back to the 1930s, prospective studies assessing its efficacy and indications remain lacking. Evidence supporting the use of surgical techniques targeting the NC is limited to small retrospective case reports and case series. A comprehensive summary of the efficacy, indications, and variations in intervention technique has yet to be reported. In the present study, we performed a systematic review of the literature published regarding the surgical intervention of the NC, providing a historical perspective for the development of NC-related interventions to the present day.

## Methods

### Literature Search and Inclusion Criteria

We conducted a systematic review according to Preferred Reporting Items for Systematic Reviews and Meta-Analyses (PRISMA) guidelines. Embase, MEDLINE, and Cochrane central databases were queried using the search strategy indicated in Supplementary Materials 1 on May 31, 2021. The search comprised of the following search terms (combination of subject headings and keywords): “caudalis”, “trigeminal nucleus”, “trigeminal tractotomy”, “trigeminal nucleotractotomy”, “trigeminal nucleotomy”, “dorsal root entry zone”, “ablation”, “lesion”, “stimulation”, and various combinations of the above.

The search strategies were modified for each database to include database-specific thesaurus terms, syntax, and field names. Only original peer-reviewed clinical studies in humans published in the English language were included. Studies were not restricted based on diagnosis or the lesioning/stimulation method. Additional relevant studies that met our inclusion criteria were identified through examination of bibliographies of relevant articles and selected reviews. Conference abstracts and unpublished studies were excluded. Anatomical studies, descriptions of technique, studies reporting on the same patient population, and reviews were excluded. Full-text analysis excluded studies with fewer than 5 patients as a method to reduce bias from case reports or small case series. The PICOS (Population, Intervention, Comparison, Outcomes, and Study Design) format was used to depict study eligibility with prespecified inclusion and exclusion criteria (Table 1).

**Table 1:**
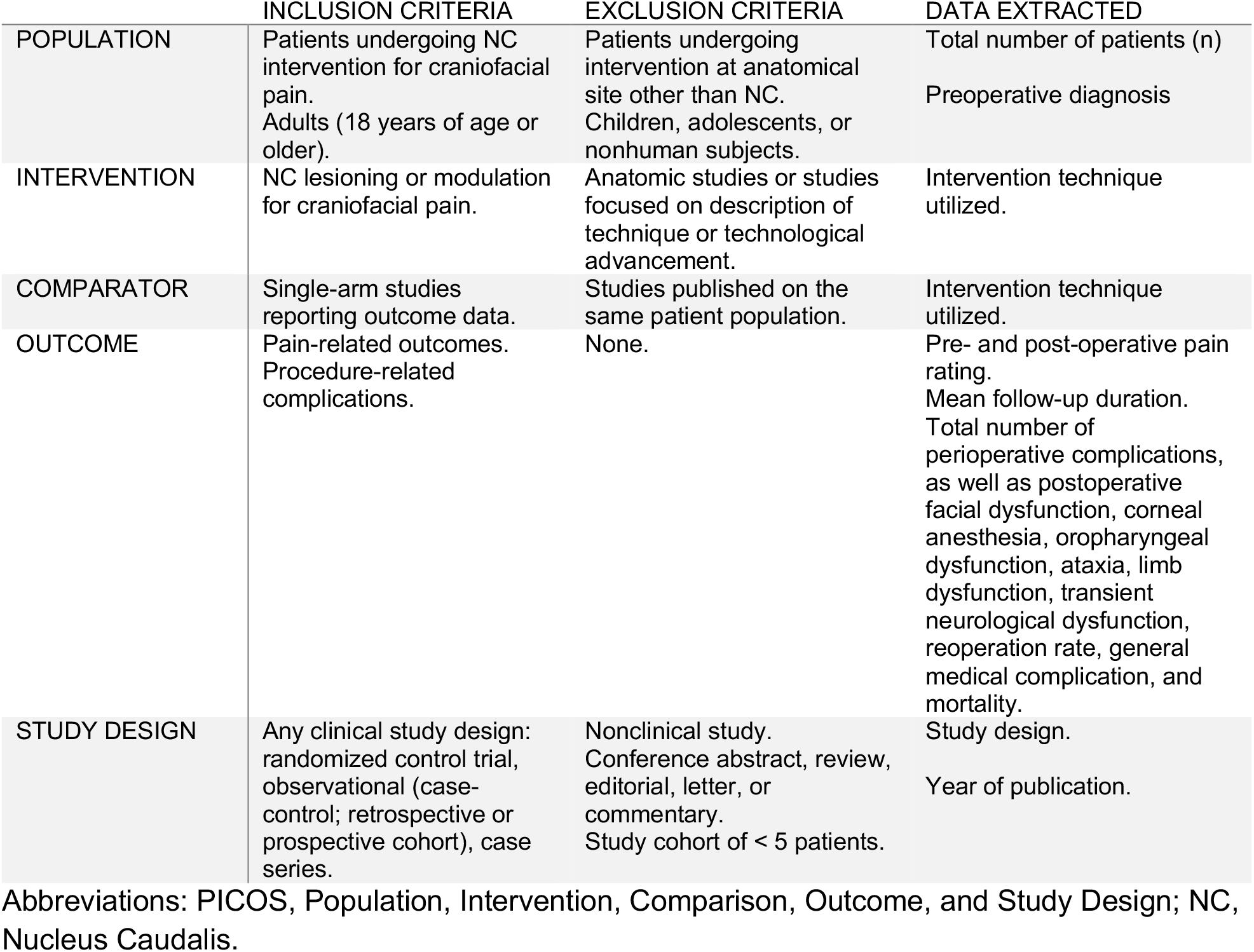
Inclusion and exclusion criteria used for article screening presented in PICOS format.

### Quality Assessment

Two reviewers (B.S., M.A.) independently screened each article title and abstract for relevance. Disagreements were resolved by discussion and consensus in the presence of a third reviewer (A.C.). All authors reviewed the preliminary results and the final analysis.

### Data Collection and Outcome Measures

Of the included studies, information regarding study type, study size, mean follow-up duration, interventional technique, preoperative diagnosis, and pre- and post-operative pain rating was collected (Table 2). Postoperative pain rating was considered a primary outcome. Mean follow-up duration was collected in years following intervention. All reported preoperative diagnoses were collected and classified based on available information. Diagnostic classification included trigeminal neuralgia or trigeminal neuropathic pain, oncologic craniofacial pain, post-herpetic pain, anesthesia dolorosa, traumatic craniofacial pain, glossopharyngeal neuralgia, post-stroke craniofacial pain, headache or migraine, multiple sclerosis-related craniofacial pain, and geniculate neuralgia. Most of the literature failed to distinguish between different classifications or causes of craniofacial pain, thereby necessitating the combination of these pathophysiological entities. Intervention techniques were classified into all procedural variations of open procedures targeting the NC or trigeminocervical complex, including nucleotomy, nucleotomy/tractotomy, and DREZ lesioning procedures; computed tomography (CT) guided and free-hand percutaneous nucleotomy/tractotomy; endoscopic DREZ nucleotomy/tractotomy; cervical spinal cord stimulation (SCS); and ultrasonic nucleotomy (Figure 1). Information regarding total perioperative complications was collected. These were further subdivided into postoperative facial dysfunction (pain, numbness, weakness), corneal anesthesia, oropharyngeal dysfunction (dysarthria, dysphagia, vocal cord paralysis), ataxia, limb dysfunction (pain, numbness, weakness, dysmetria), transient dysfunction (pain, numbness, weakness, ataxia), reoperation rate, general medical complication, and mortality.

**Table 2:**
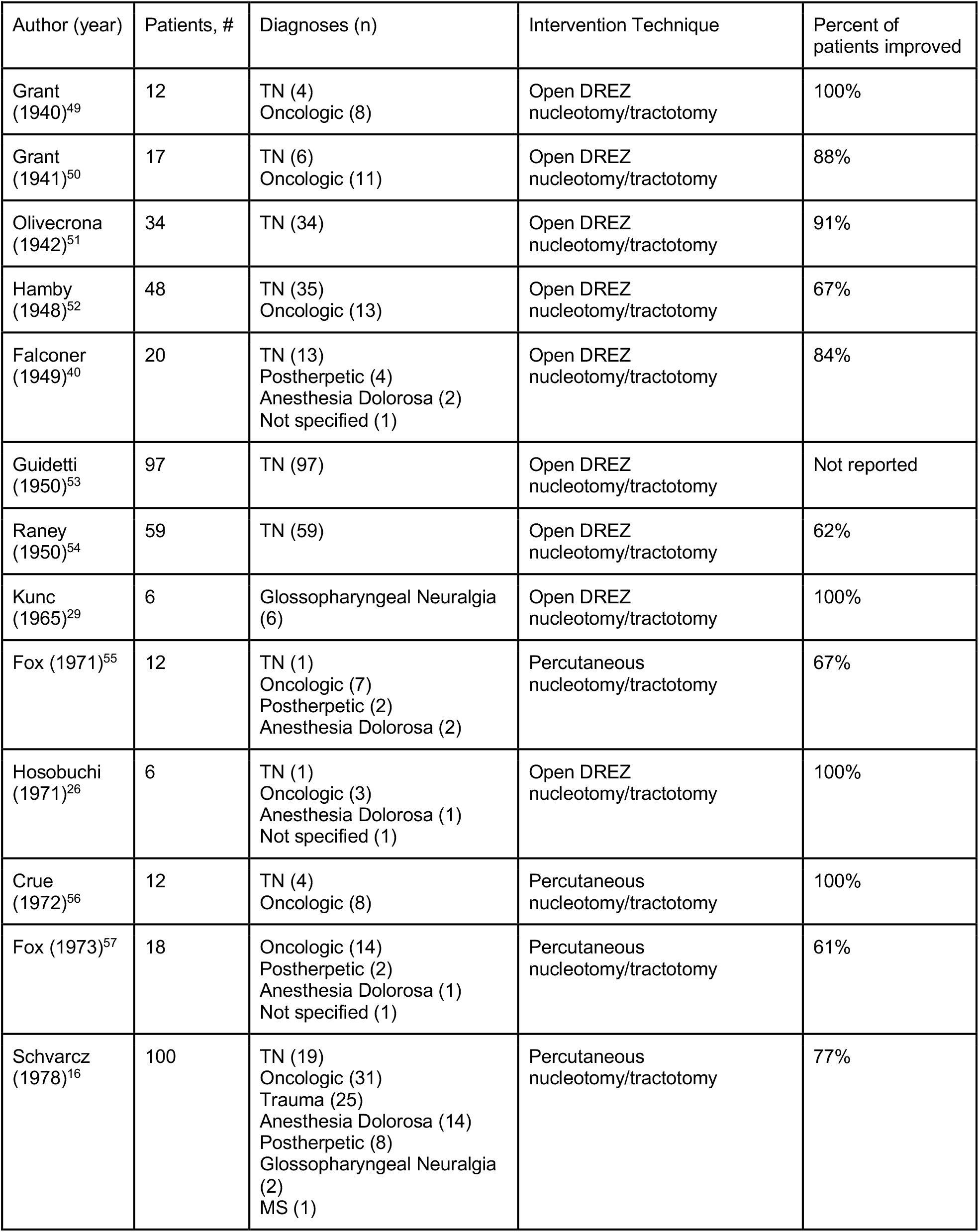
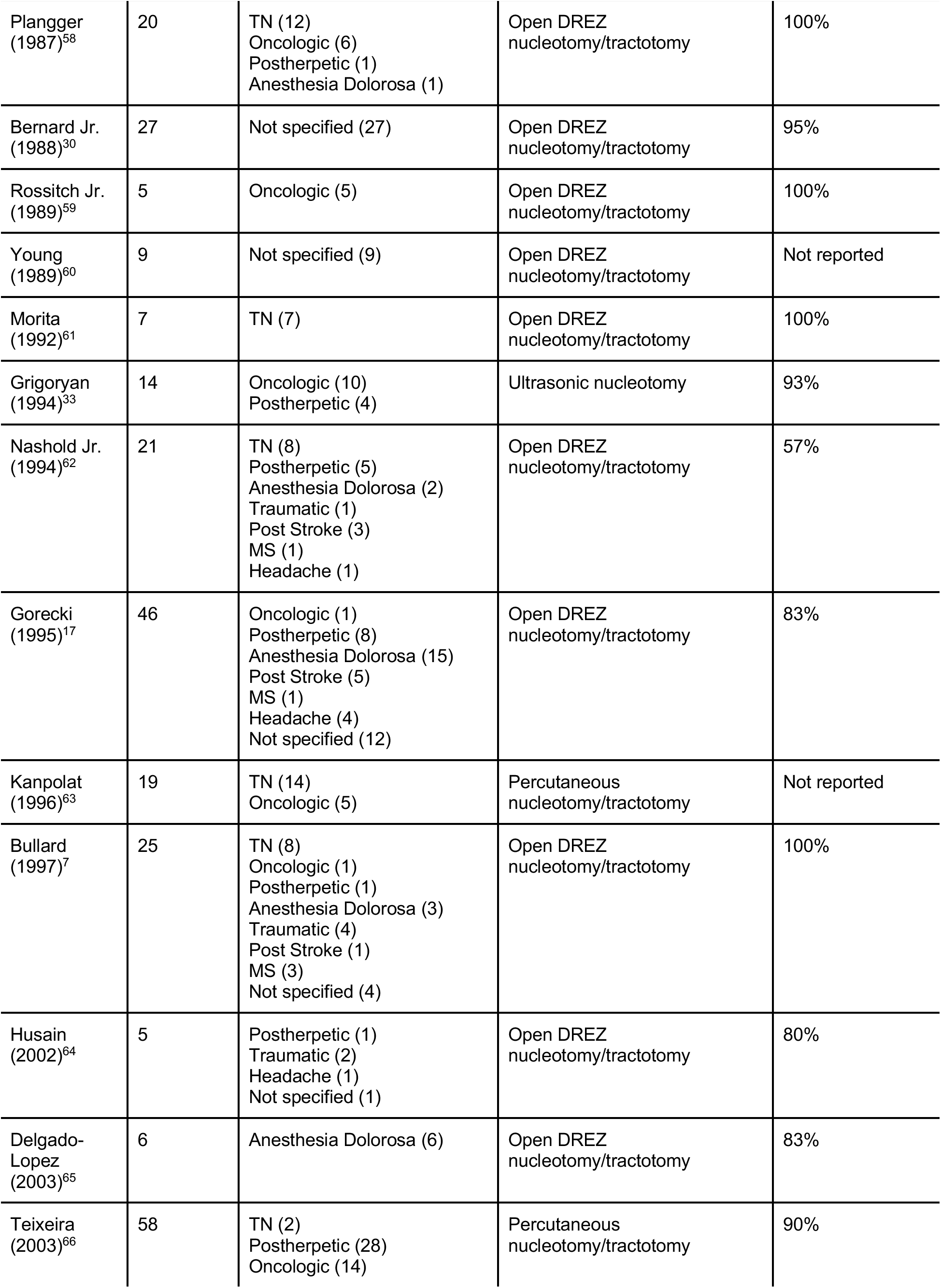
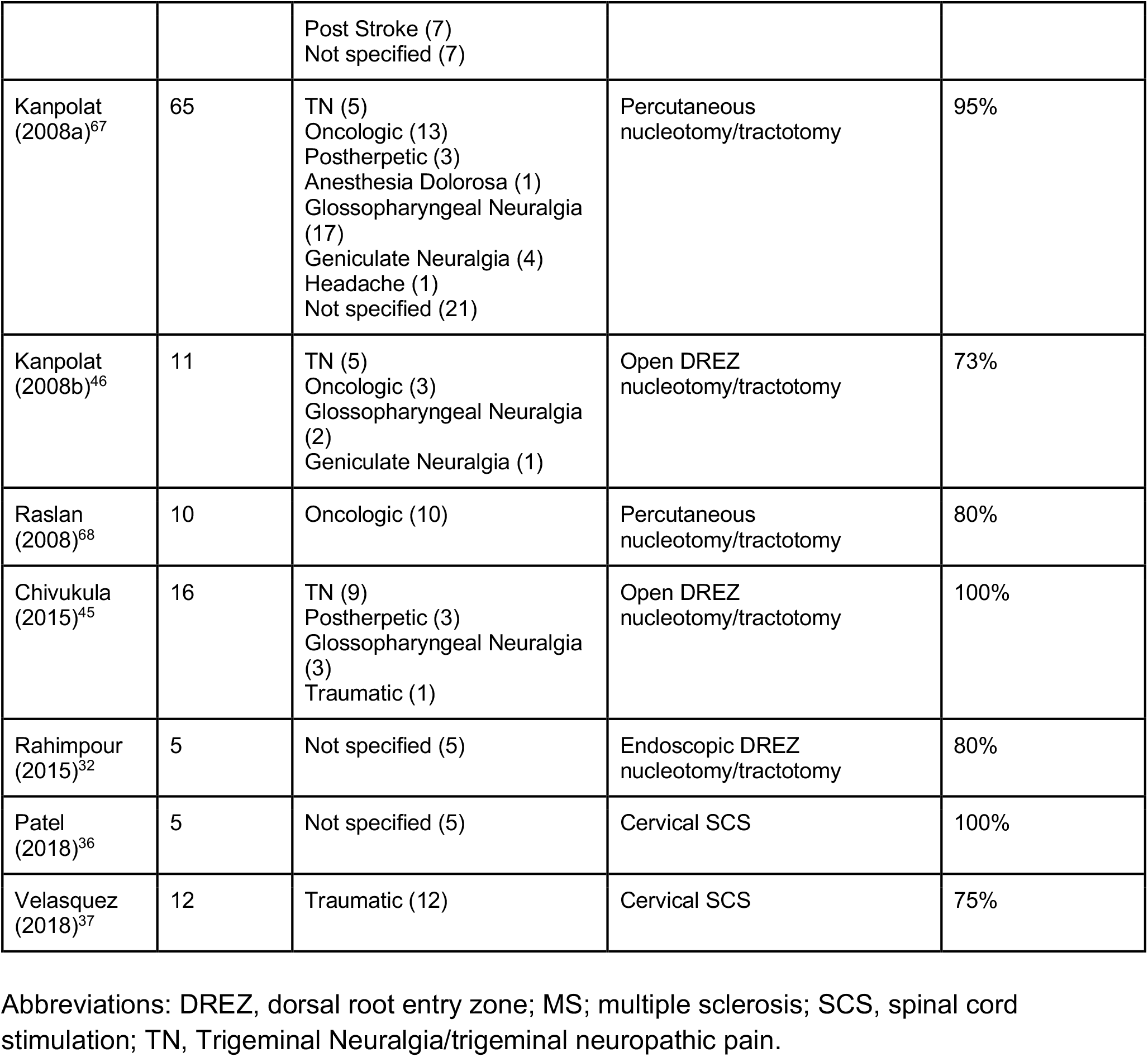
Summary of articles reporting interventions targeting the nucleus caudalis for craniofacial pain.

**Figure 1:**
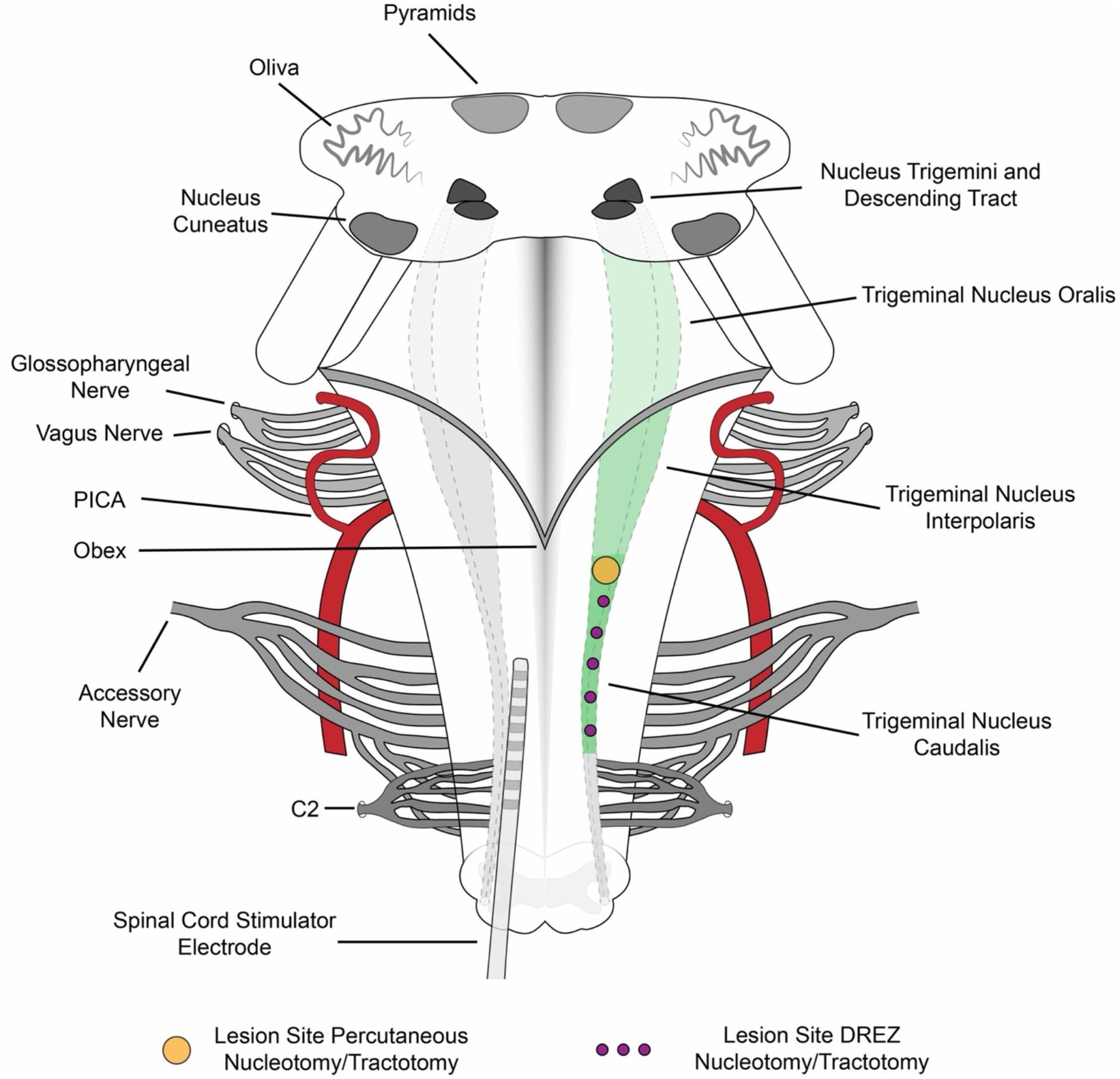
Illustration of the dorsal aspect of the cervicomedullary junction, depicting the lesion site for nucleus caudalis interventions including DREZ nucleotomy/tractotomy, percutaneous nucleotomy/tractotomy, and cervical SCS. Abbreviations: DREZ, dorsal root entry zone; SCS, spinal cord stimulation.

Due to the historical nature of this review spanning back to the first descriptions of interventional procedures involving the NC, we observed a wide variability of methods for reporting pain in the literature. Most commonly, qualitative and non-standardized methods were used to report pain outcomes with only 5 studies reporting using the standardized Visual Analog Scale. To accommodate this and facilitate the comparison of modern and classic studies, a composite rating scale was employed in this review (Table 3). A composite pain score of 1 includes patients that are pain-free, Visual Analog Scale (VAS) 0, or have had a 100% pain reduction. A pain score of 2 includes those with a “good” response, VAS 1-3, or a >50% pain reduction. A pain score of 3 includes those with a “satisfactory” or “fair” response, VAS 4-6, or a <50% pain reduction. Finally, a pain score of 4 includes those who experienced no improvement, pain worsening, or VAS 7-10.

**Table 3:** Composite pain reporting scale.

| Composite pain score | Description |
| --- | --- |
| 1 | Pain free, VAS 0, 100% pain reduction |
| 2 | “Good” response, VAS 1-3, >50% pain reduction |
| 3 | “Satisfactory” or “fair” response, VAS 4-6, <50% pain reduction |
| 4 | No improvement, pain worsening, VAS 7-10 |

Abbreviations: VAS, visual analog scale.

The fraction of patients demonstrating any symptomatic improvement post-procedure (pain score of 1-3) was calculated as a percentage of the total number of subjects included in the study (Table 2). Patients with mortality prior to follow-up or if the outcome was not reported were deemed outcome unknown. To determine the relative surgical success between techniques, they were divided as follows: open focal lesioning techniques (nucleotomy/tractotomy), DREZ (multipoint lesioning technique), percutaneous nucleotomy/tractotomy, and neuromodulation (cervical SCS). Pain outcome distribution was compared between each using a Chi-Squared test. The fraction of patients with post-procedural improvement was assessed with respect to publication date to determine the influence of procedural modification over time (Supplementary Materials 3). This analysis used a linear model for all intervention types as well as individually for those with an inadequate number of studies. Linear models were implemented using the R statistical language and environment.^20^ Event rates for composite scores and complications were pooled across studies, and logit transformed with 95% confidence intervals calculated assuming a binomial distribution. This was calculated using MATLAB R2019b.^21^

## Results

A total of 782 articles were identified through the literature search and underwent title and abstract review (Figure 2). Two separate reviewers (B.S. and M.A.) screened each article to meet inclusion/exclusion criteria. Following removal of duplicates, review of bibliographies from relevant articles, and exclusion of studies with n < 5 patients (Supplementary Materials 2), 33 studies encompassing 827 patients were included for analysis as summarized in Table 2. Disagreement was resolved by a third reviewer (A.C.). All identified studies for inclusion were case series published between 1940 and 2020. No randomized or prospective trials were identified. The results of the data extraction for included studies are summarized in Table 2. The publications included in this work represent an 80-year history of techniques targeting the NC for treatment of craniofacial pain (Figure 3). The rate of publications continues to increase over time and there are emerging techniques including endoscopic approaches and spinal cord stimulation (SCS) seen in the last 10 years (Figure 3)

**Figure 2:**
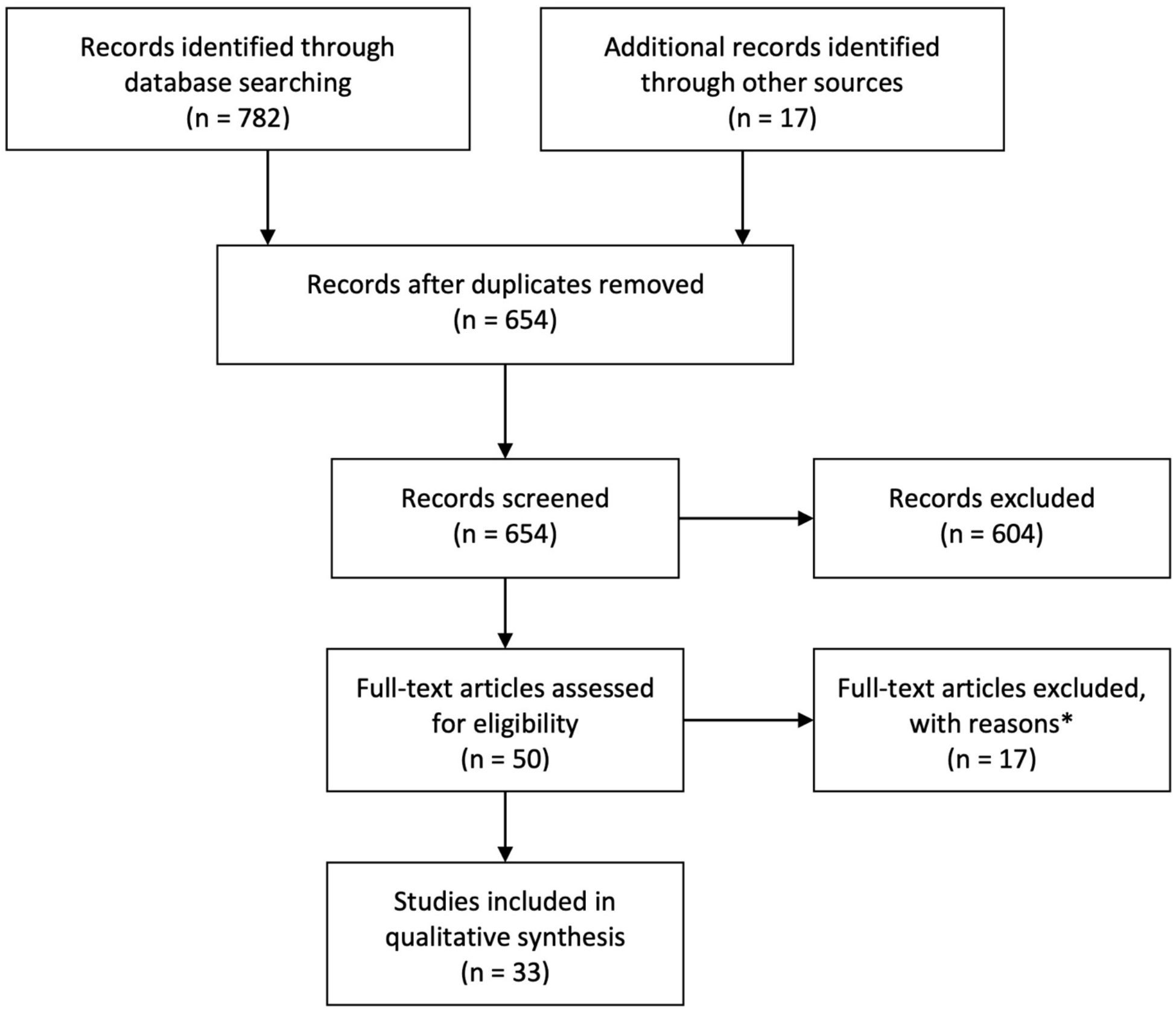
Preferred Reporting Items for Systematic Reviews and Meta-Analyses (PRISMA) flowchart outlining article selection. *Articles with fewer than 5 patients (Supplementary Materials 2).

**Figure 3:**
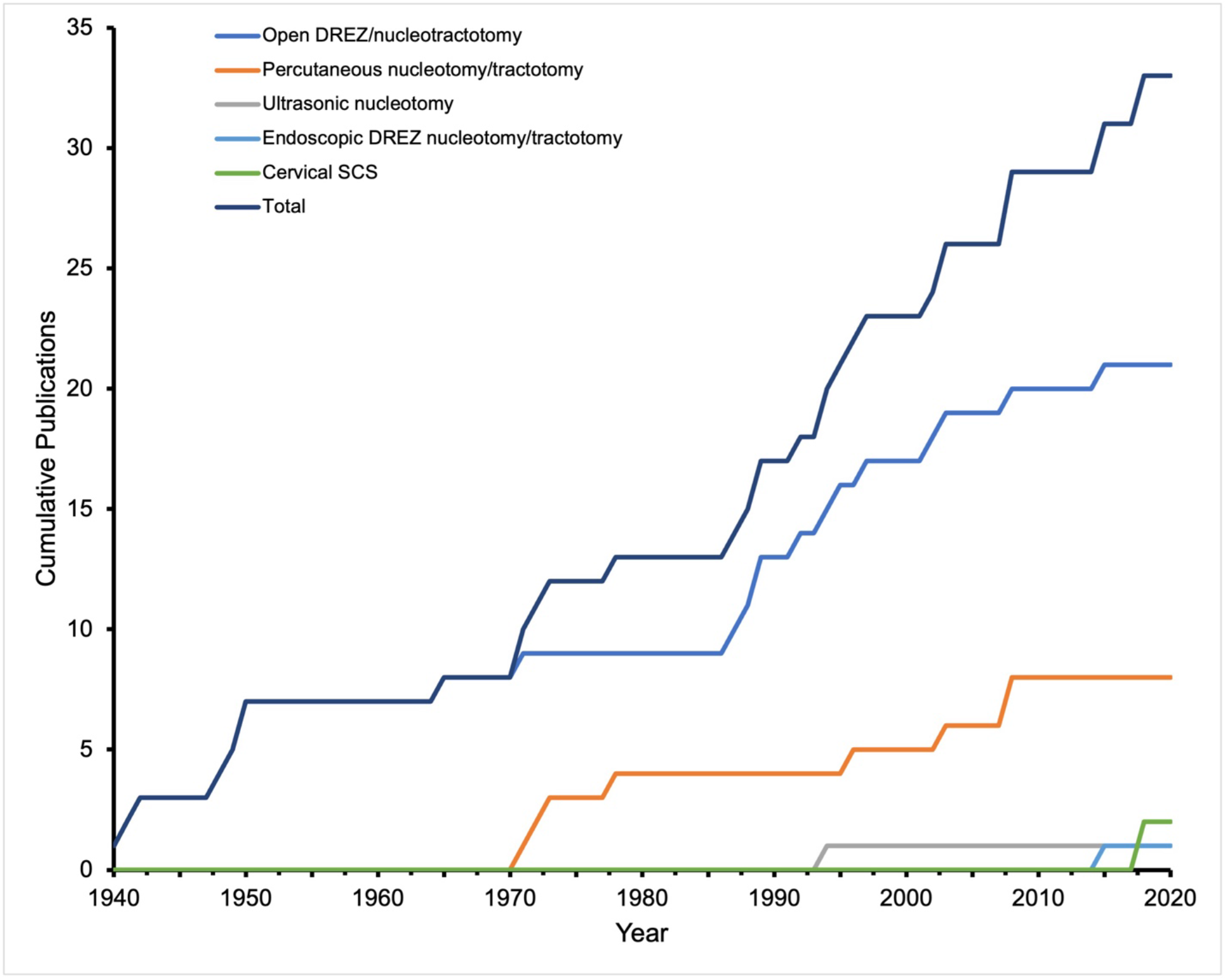
Cumulative number of studies examining nucleus caudalis targeting interventions in the treatment of craniofacial pain. Over the last 80 years, 33 articles have been published investigating techniques targeting the NC for treatment of craniofacial pain. The 1940s and 1970s saw the development of open (blue) and percutaneous approaches (orange) respectively. The rate of publication continues to increase over time and novel techniques continue to emerge including endoscopic approaches (light blue) and SCS (green) seen in the last 10 years and ultrasonic (grey) previously. Abbreviations: DREZ, dorsal root entry zone; NC, nucleus caudalis; SCS, spinal cord stimulation.

### Preoperative Diagnosis

Treatments targeting the nucleus caudalis have been used to ameliorate craniofacial pain in a wide range of patient populations. Of the 827 patients included in this review, 41.5% (n=343) had a diagnosis of trigeminal neuralgia or trigeminal neuropathic pain (Figure 4). Oncologic craniofacial pain comprised 19.7% (n=163), 8.5% (n=70) had post herpetic pain, 5.7% (n=47) had anesthesia dolorosa, 5.6% (n=46) had traumatic craniofacial pain, 3.6% (n=30) had glossopharyngeal neuralgia, 1.9% (n=16) had post-stroke craniofacial pain, 0.8% (n=7) had headache or migraine, 0.7% (n=6) had multiple sclerosis related craniofacial pain, 0.6% (n=5) had geniculate neuralgia, and 11.4% (n=94) had multiple diagnoses or unspecified craniofacial pain (Figure 4).

**Figure 4:**
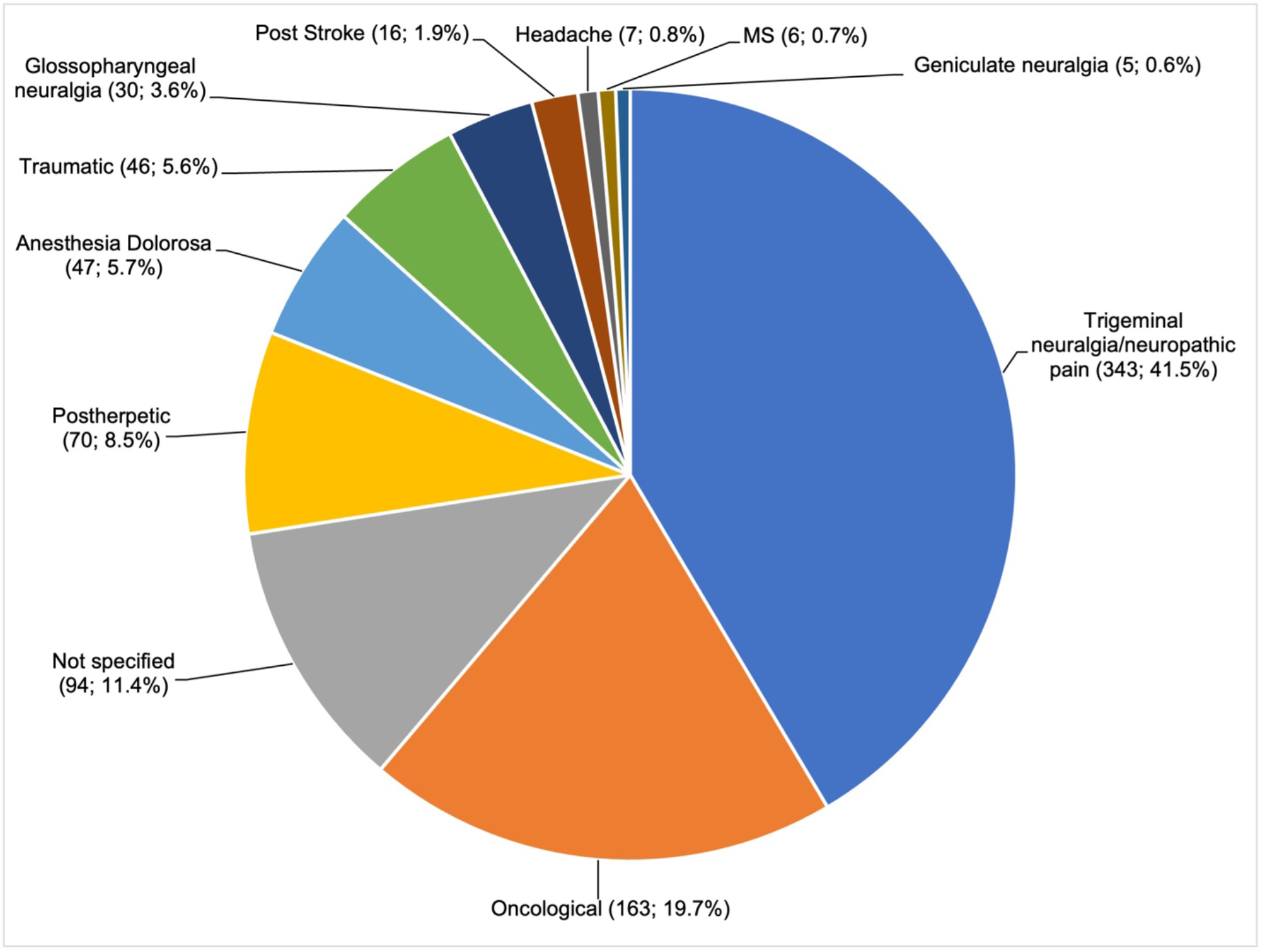
Distribution of preoperative diagnoses. Thirty-three articles reporting on 827 patients demonstrate a wide range of sources of craniofacial pain treated with NC interventions. The most common being trigeminal neuralgia/trigeminal neuropathic pain (n=343, 41.5%), oncological (n=163, 19.7%), and postherpetic pain (n=70, 8.5%). Rarer causes of craniofacial pain are also represented, such as headache (n=7, 0.8%), MS (n=6, 0.7%), and geniculate neuralgia (n=5, 0.6%). Abbreviations: MS, multiple sclerosis; NC, nucleus caudalis.

### Intervention Technique

Review of the included articles revealed 5 intervention techniques targeting the nucleus caudalis for craniofacial pain (Figures 1 and 5). The most common approach was open NC DREZ nucleotomy/tractotomy, reported in 21 studies and comprising 60.1% of patients (n=497). CT guided and free-hand percutaneous nucleotomy/tractotomy is reported in 8 studies and 35.6% of patients (n=294). Cervical SCS is reported in 2 studies and 2.1% of patients (n=17). Ultrasonic nucleotomy is reported in one study and 1.7% of patients (n=14). Endoscopic DREZ nucleotomy/tractotomy is reported in 1 study and 0.6% of patients (n=5). Although not included in the analysis, Gamma Knife radiosurgery was reported in a single case series of 2 patients.^22^

**Figure 5:**
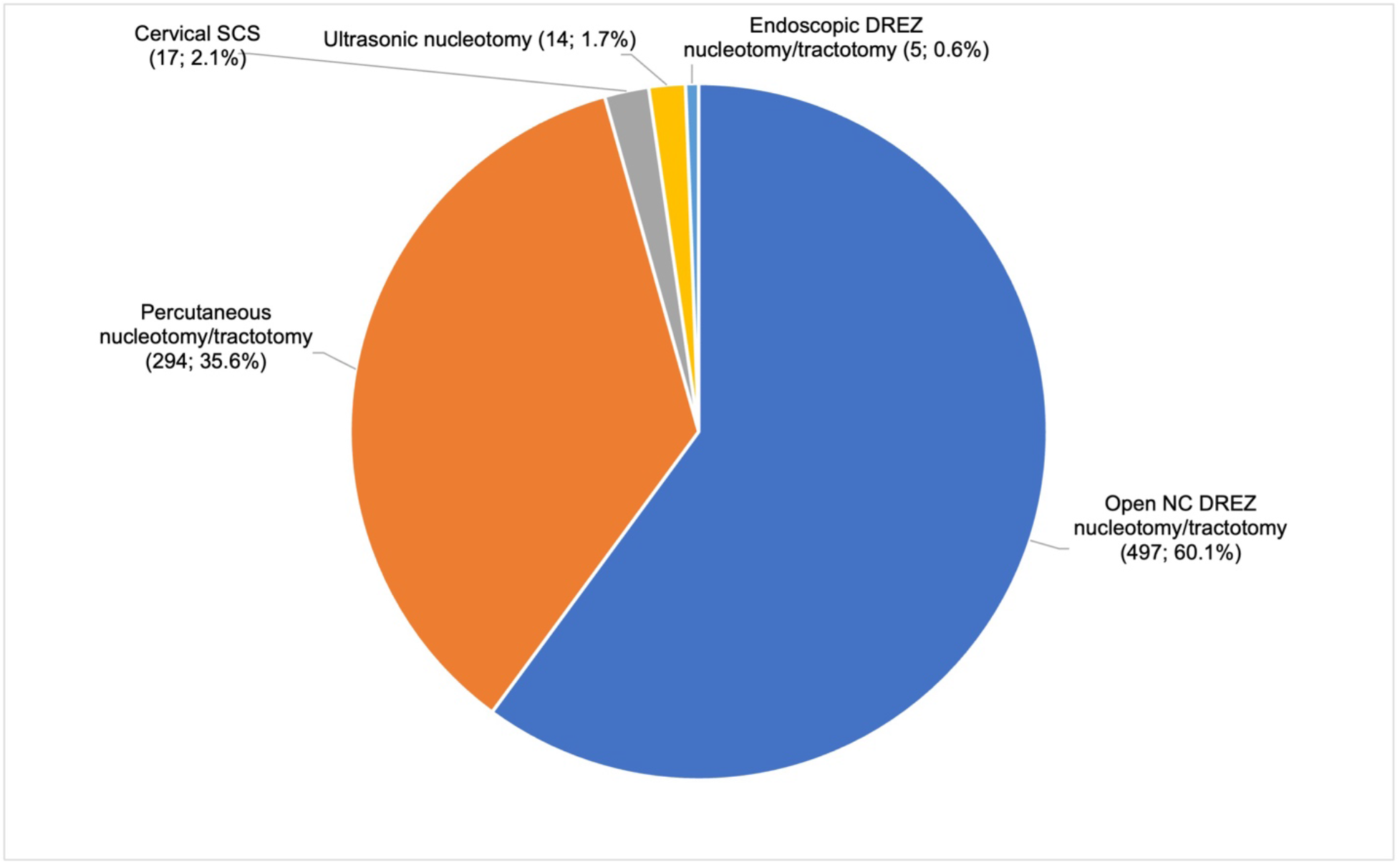
Distribution of intervention technique. The included studies encompass 5 main intervention techniques targeting the NC. In order of decreasing prevalence: Open NC DREZ nucleotomy/tractotomy (n=497, 60.1%), Percutaneous nucleotomy/tractotomy (n=294, 35.6%), Cervical SCS (n=17, 2.1%), Ultrasonic nucleotomy (n=14, 1.7%), and Endoscopic DREZ nucleotomy/tractotomy (n=5, 0.6). Abbreviations: DREZ, dorsal root entry zone; NC, nucleus caudalis; SCS, spinal cord stimulation.

### Treatment Outcomes

As mentioned previously, various pain reporting methods are utilized in the studies reviewed. To facilitate comparison of treatment outcomes between studies, a composite pain rating scale was developed and outlined in Table 3. Preoperative pain is even less consistently reported. Of the 827 included patients, 142 (17.2%) have preoperative VAS reported with a mean of 8.6. Using the implemented composite score developed to facilitate comparison between modern and historical studies, interventions targeting the NC resulted in a pain score of 1 (i.e., pain freedom, VAS 0, or 100% pain reduction) in 237 patients (28.7%), pain score of 2 (i.e., “good” response, VAS 1-3, or a >50% pain reduction) in 215 patients (26.0%), pain score of 3 (i.e., “satisfactory” or “fair” response, VAS 4-6, or a <50% pain reduction) in 35 patients (4.2%), pain score of 4 (i.e., no improvement, pain worsening, or VAS 7-10) in 49 patients (5.7%) (Figure 6). The treatment outcome was unknown in 291 patients (35.2%). This includes patients lost to follow-up, passed away, or inadequately reported.

**Figure 6:**
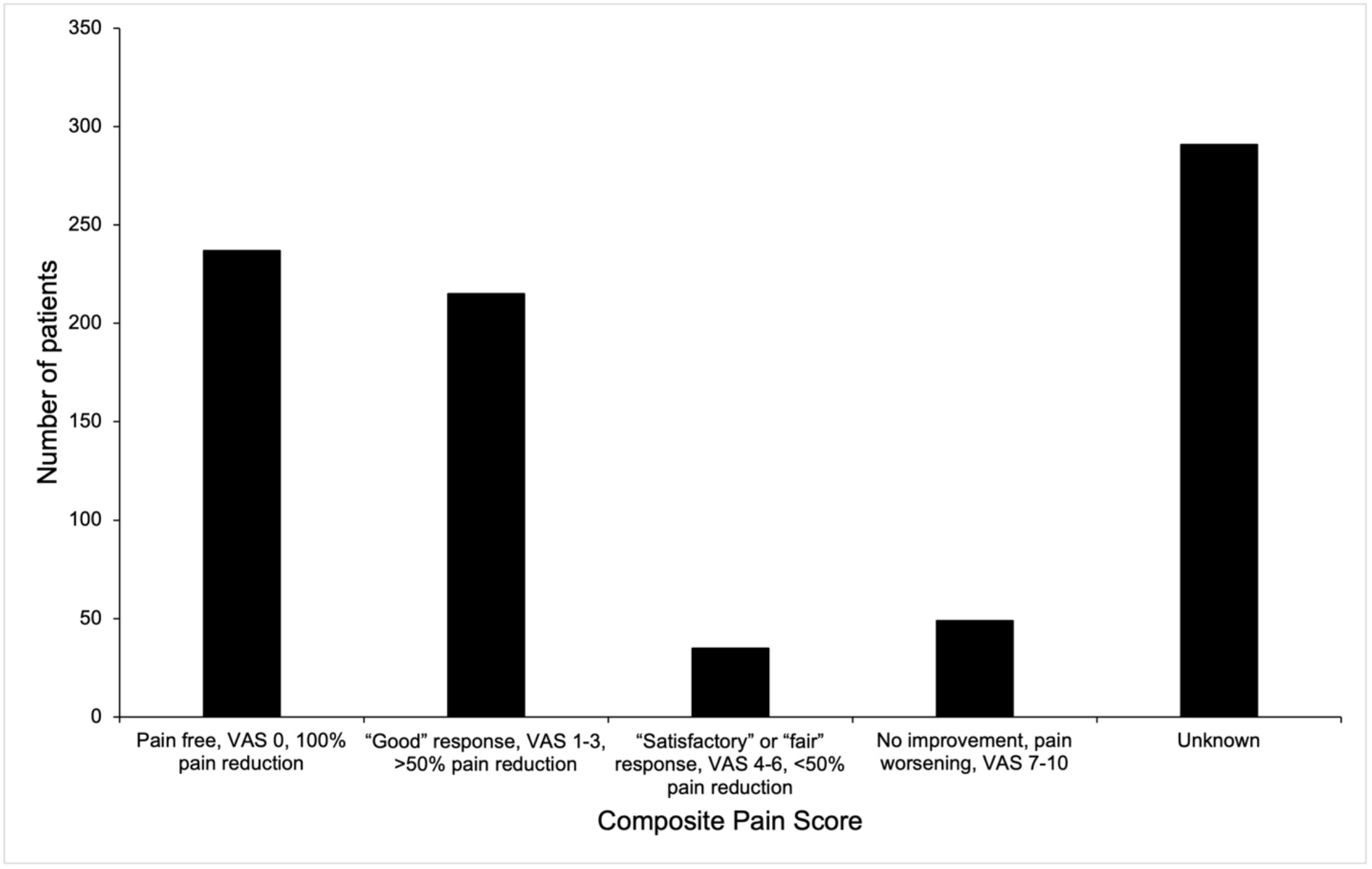
Post-intervention craniofacial pain outcome. Intervention efficacy was reported for 536 patients. A composite pain score was created to facilitate comparison between modern and historical studies. Of those with reported outcome 44.2% (n=237) were pain free, 40.1% (n=215) had a “good” response, 6.5% (n=35) had a “satisfactory” response, and 9.1% (n=49) had no improvement or worsening pain. Abbreviation: VAS, visual analog scale.

Focusing on subjects with clearly reported outcome values, the intervention types were reclassified as open focal lesioning techniques (nucleotomy/tractotomy) (n patients = 154), open DREZ (multipoint lesioning technique) (n patients = 141), percutaneous nucleotomy/tractotomy (n patients = 224), and cervical SCS (n patients = 17) and outcome distribution were compared between the types (Figure 7). Chi-squared demonstrates a significant difference in the outcome distribution between the intervention types (χ ^2^ = 57.5322, p <0.001) (Supplemental Material 3). Of the 22 studies (66.7%) that reported follow up duration, the mean duration was 2.44 years. When stratified by intervention type, the mean follow-up duration for open focal lesioning techniques (nucleotomy/tractotomy) was 1.66 years, open DREZ (multipoint lesioning technique) was 2.46 years, percutaneous nucleotomy/tractotomy was 2.10 years, and for cervical SCS was 3.55 years.

**Figure 7:**
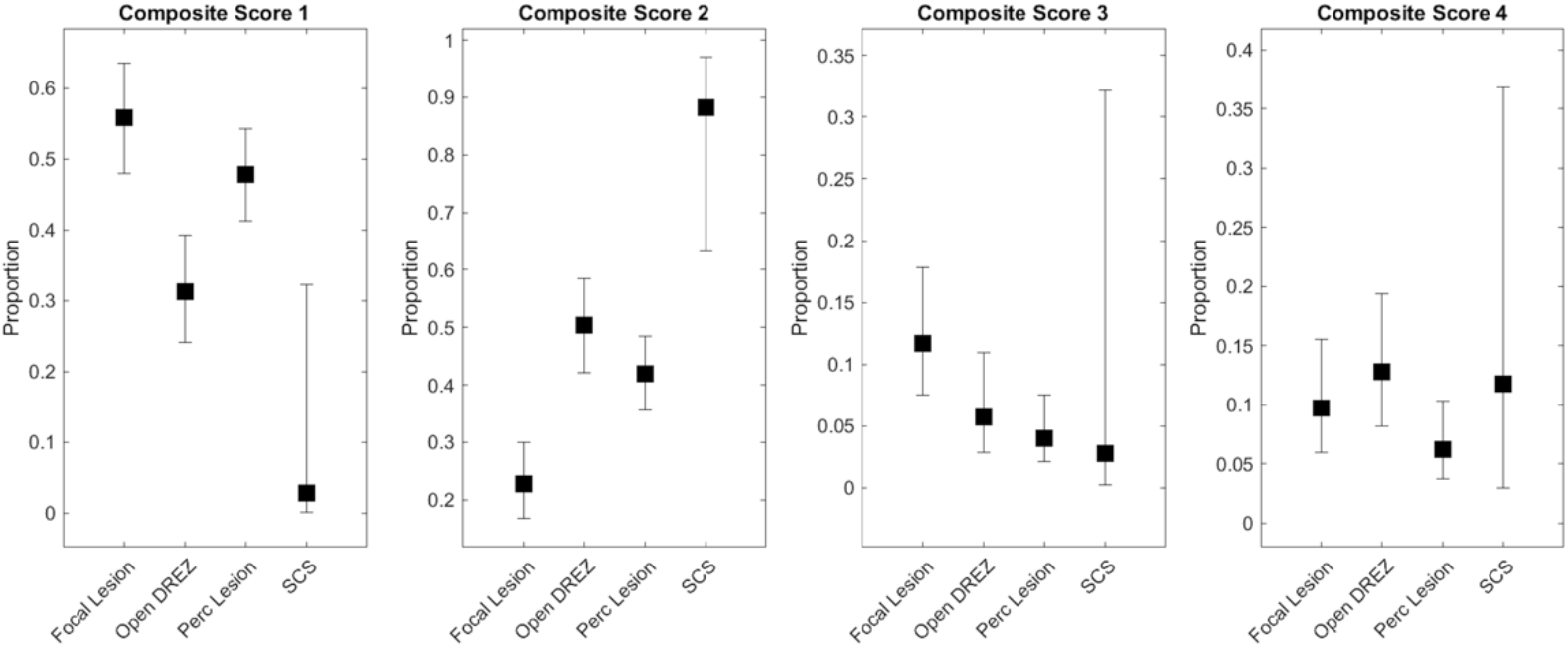
Distribution of pain outcomes by intervention technique. The intervention types were reclassified as open focal lesioning techniques (nucleotomy/tractotomy) (n patients = 154), open DREZ (multipoint lesioning technique) (n patients = 141), percutaneous nucleotomy/tractotomy (n patients = 224), and cervical SCS (n patients = 17). Proportion of patients with a given outcome was reported with error bars representing 95% confidence intervals. Chi-squared demonstrates a significant difference in the outcome distribution between the intervention types (χ ^2^ = 57.5322, p <0.001). Abbreviations: DREZ, dorsal root entry zone; Perc, percutaneous; SCS, spinal cord stimulation.

Linear regression failed to demonstrate a trend toward improving postoperative outcomes in interventions over the 80-year history (Supplemental Materials 4). A non-significant trend towards improved outcomes is seen in open focal lesioning and percutaneous nucleotomy/tractotomy when separated by intervention type. The correlation coefficients are 0.45 and 0.44, the covariances are 1.05 and 1.12, and p-values are 0.19 and 0.33 respectively (Supplemental Materials 4).

### Complications

A total of 251 complications are reported. The proportion of individual complications per intervention type is presented in Figure 8. All complications except corneal anesthesia (n = 5), oropharyngeal dysfunction (n = 11), and limb dysfunction (n = 14) demonstrate a statistically significant difference in distribution between intervention types (focal lesioning techniques, DREZ, percutaneous, and neuromodulation) (p < 0.001) (Supplemental Material 5).

**Figure 8:**
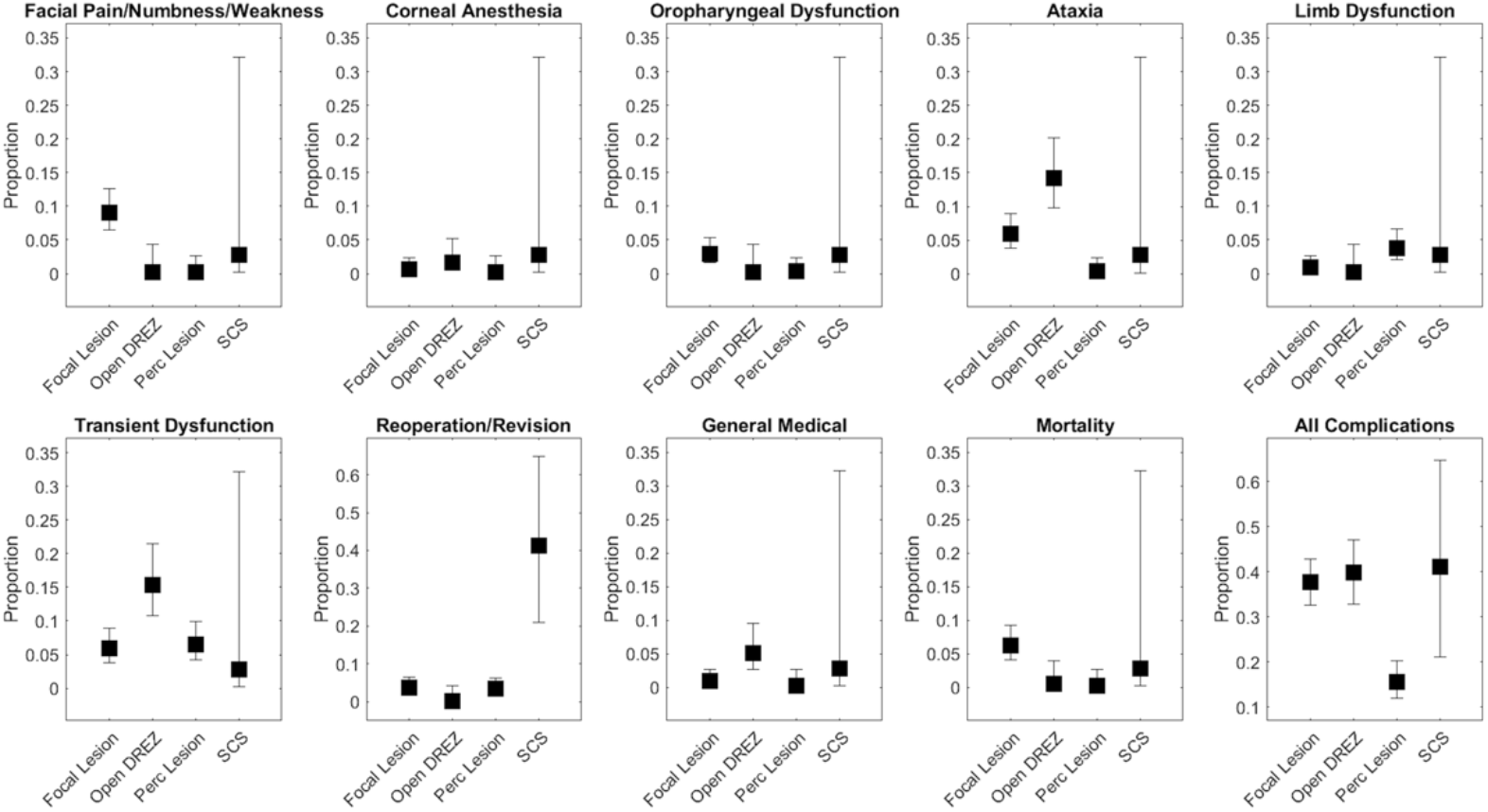
Proportion of patients with a given complication arranged by intervention technique. All complications except corneal anesthesia (n = 5), oropharyngeal dysfunction (n = 11), and limb dysfunction (n = 14) demonstrate a statistically significant difference in distribution between intervention types. Proportion of patients with a given complication is reported with error bars representing 95% confidence intervals. See Supplemental Material 5 for data and statistical results. Abbreviations: DREZ, dorsal root entry zone; Perc, percutaneous; SCS, spinal cord stimulation.

## Discussion

In this study, we performed a systematic review of the published literature regarding the surgical interventions of the NC and investigated the efficacy and indications for these procedures. 87% of subjects demonstrated some improvement from NC-targeted treatment. Furthermore, 44.2% reported complete pain freedom postoperatively. We show percutaneous lesioning has a relatively lower complication rate compared to other intervention types. Despite the increase in described surgical options for craniofacial pain, up to 27% of patients undergoing surgical treatment have insufficiently managed pain at 5 years.^2,23^ The present study demonstrates that there has been a recent increase in publication rate over 80 years since the original descriptions of nucleus caudalis interventions. Contributing to this are not only the advancements in open and percutaneous techniques but also the emergence of new approaches such as ultrasonic lesioning, radiosurgery, and SCS.

The studies included in this review report on diverse diagnoses resulting in chronic craniofacial pain including post-herpetic pain, trigeminal neuralgia or trigeminal neuropathic pain, oncological pain, migraine, multiple sclerosis-related craniofacial pain, and geniculate neuralgia. A detailed description regarding the pathophysiology of pain associated with each of these conditions is outside the scope of this review; however, common to each of these diseases is injury (compression, denervation/deafferentation, demyelination) of the general somatic afferent fibers of the Vth, VIIth, IXth, or Xth cranial nerves. These fibers synapse onto second order neurons within the descending trigeminocervical complex, which extends from the pons to the upper cervical spinal cord on the dorsal surface of the brainstem.^24^ Based on anatomic and clinical data, the nucleus caudalis was identified as the most important locus for modulation, integration, and conduction of nociception from craniofacial structures to higher-order intracranial structures for pain perception.^9,25–28^ This provides a logical target for treatment of pain associated with injury to cranial nerves V, VII, IX, and X. Intervention techniques such as the NC DREZ nucleotomy/tractotomy utilize a small suboccipital craniectomy and C1-C2 laminectomy to elevate the cerebellar tonsils and visualize the obex. The NC occupies the triangular area between the dorsolateral sulcus and the emerging points of the accessory nerve. It tapers caudally before joining the spinal DREZ of C2. During NC DREZ nucleotomy/tractotomy multiple rostrocaudal and mediolateral lesions are made at a depth of 3-4mm covering 5mm below the obex to the upper dorsal rootlets of C2.^24^

The identified articles reveal progressive adaptation of intervention techniques targeting the NC. Most studies (21 studies comprising 60.1% of patients) report on the open NC DREZ nucleotomytractotomy. Open procedures in this region date back to the 1930s where Sjoqvist demonstrated that sectioning of the trigeminocervical complex at the level of the medulla above the obex resulted in ipsilateral thermoanalgesia of the face.^9^ Although initially plagued by complications, a near century of surgical advancement has considerably improved its safety and efficacy, including the development of the more selective vertical trigeminal partial nucleotomy in 1965.^29^ The 1970s saw the emergence of stereotactic and percutaneous targeting techniques, improving pain outcomes through patient cooperation and intraoperative monitoring of lesion-induced analgesia under local anesthetic.^15^ Surgical refinement and improved anatomical understanding of the region in the 1970s and 80s resulted in specific open NC DREZ lesioning procedures gaining popularity with higher immediate success rates.^30^ In recent decades a trend towards less invasive techniques is apparent, as morbidity related to injury of nearby structures of the spinal cord and medulla from open procedures remain high; historically 5% encounter ataxia, 25% with paresthesias, and 8-24% with lesioning of the 10^th^ nerve.^24^ Endoscopic NC DREZ nucleotomy/tractotomy procedures have been developed to minimize the morbidity associated with large incisions and craniotomies.^31,32^ Noninvasive techniques using ultrasound^33^ and Gamma Knife radiosurgery^22^ have similarly been developed. Finally, the most recent adaptation of techniques targeting the NC is in the form of spinal cord stimulation as a titratable and reversible form of neuromodulation of the region.^34–38^

It is clear that the techniques targeting the NC for ablation or neuromodulation represent an additional option for the treatment of intractable craniofacial pain of diverse pathophysiological origins. Despite the variations in intervention technique and preoperative diagnosis, 87% of subjects demonstrated some improvement from treatment. Furthermore, 44.2% reported complete pain freedom postoperatively, suggesting remarkable efficacy for a group of treatments typically reserved for complex and otherwise surgically and medically refractory craniofacial pain. Although this might be an overestimation of the overall effectiveness of these procedures due to publication bias, it is nevertheless encouraging for continued prospective study of these procedures. Also demonstrated here are significant differences in outcome distributions between different procedural techniques. However, due to the inconsistencies in outcome reporting from the inclusion of studies prior to standardization of reporting pain metrics, efficacy comparisons between intervention type or preoperative diagnosis could not be reliably made. There have been efforts in the literature to standardize pain outcome reporting, with multiple well-validated pain rating scales available.^39^ Here we present added evidence for the need to continually report outcomes in a way that allows for robust comparisons. It is imperative that future prospective studies address these lingering questions.

Analysis of reported complication rates demonstrates significant differences in complications between the different intervention techniques. The data support percutaneous interventions having a relatively lower rate of complications compared to others (Figure 8). These results should be interpreted cautiously as they are certainly confounded by publication bias, inadequate reporting of complications, limited duration of follow-up (2.44 years), and lack of longitudinal outcome reporting. However, encouragingly, a treatment response durability of at least a year is demonstrated for each intervention type. Further investigation should be the subject of future prospective trials.

Open trigeminal tractotomy (focal lesioning technique) as a procedure has become largely historical due to frequent partial/patchy craniofacial analgesia^40^, high recurrence rates^41^, and frequency of morbidity and mortality.^24^ Advancements in technology and understanding in the 1970s paved the way for the more common adaptation of these procedures seen today: caudalis DREZ lesioning (multipoint lesioning technique) and percutaneous lesioning. Determining when to pursue each technique and for whom remains a difficult question to answer when prospective trials and comparison studies are lacking. The literature reviewed here suggests that with caudalis DREZ lesioning pain improvement may be achieved in 70% patients. Bernard et al. surmise that patient description of pain may help predict postoperative outcome^30^, patients with sharp/burning pain finding better relief than those with dull pain, 55% and 24% respectively.^30^ Better results were also achieved when pain is restricted to the trigeminal territory and with fewer trigeminal distributions involved.^30^ Good outcome is observed in 75% of patients with pain involving one trigeminal distribution, 50% of patients with two, and 38% of patients where all three territories are involved.^30^ Similarly percutaneous procedures demonstrate pain improvement in 71% of patients. However, these procedures as described in the literature tend to target afferent fiber tracts, second-order neurons, and internuncial neurons and tracts in the spinal trigeminal complex eliminating abnormal discharging of deafferented neurons and thereby providing better pain relief in patients with central or deafferentation pain compared to caudalis DREZ lesioning.^18,30,33,42,43^ Furthermore, we demonstrate a lower total complication rate in percutaneous techniques with only a slightly higher reoperation/revision rate compared to caudalis DREZ lesions. Historical literature and the results from this work support the use of percutaneous procedures in providing an equivalent degree of craniofacial pain reduction as caudalis DREZ lesioning, with better efficacy in deafferentation pain, and fewer complications. Caudalis DREZ lesioning may be reserved for uncooperative patients or those with pain refractory to other treatments, and those with sharp/burning pain in a limited distribution.

Newer techniques such as endoscopic, ultrasound, gamma knife, and SCS report on too few patients to make meaningful conclusions regarding their indications and relative efficacy, however neuromodulation with cervical SCS is showing a recent rise in publication rate.^34–38^ The efficacy and safety of SCS in the treatment of chronic pain is well accepted and supported in the literature.^44^ Likewise, this efficacy is replicated in this review, where 88% of patients receiving cervical SCS experience some degree of pain reduction. Velasquez et al. report a mean pain reduction of 57.1% making it comparable to ablative procedures on a patient level.^37,45,46^ However large prospective studies are lacking and indication recommendations are limited. We show a lower overall complication rate relative to caudalis DREZ lesioning with no reported mortality or permanent neurological deficit such as ataxia, weakness, or paresthesia. There was a slightly higher total complication rate compared to percutaneous techniques, likely secondary to a higher reoperation/revision rate from infection, loss of efficacy, and lead malpositioning rather than neurological deficit.^37^ With a body of literature that is continually growing, cervical SCS is presenting itself as a potential early, titratable, and reversible therapy with a lower complication risk and comparable outcomes.

The need for ongoing innovation in chronic pain management is apparent and less invasive methods are emerging. Of particular interest, recent reports have described both intraventricular and intrathecal analgesia administration in cases of refractory craniofacial pain.^47,48^ As the evidence continues to evolve these procedures may become viable options for patients that have failed other strategies or whose pain is bilateral.^47,48^

The first application of lesioning of the trigeminocervical complex dates back to the 1930s^9^; however, evidence supporting its use is limited to small retrospective case series and case reports. These study designs are particularly at risk of several biases potentially confounding the results presented here, including selection bias, recording bias, misclassification bias, and uncontrolled confounding bias. Furthermore, as mentioned above the limitations encountered are a lack of prospective studies and inadequate outcome reporting to strengthen comparisons between intervention types and diagnosis. These limitations have restricted the critical appraisal of the sources of evidence for risk of bias assessment. Historical studies, as encountered in this work, particularly suffer from a lack of access to more recently devised outcome reporting schemes and classification systems. For example, the contemporary classification of trigeminal neuralgia and as such differentiating trigeminal neuropathic pain from trigeminal neuralgia could not be performed. Furthermore, the heterogeneity of included studies with respect to intervention, diagnosis, and outcome reporting measures necessitated grouping all these factors together and creating a custom post-procedural pain reporting scale. This may limit the strength of efficacy conclusions; however, it does contribute greatly to the generalizability of the included results and allowed comparison to historical studies. Additionally, information regarding follow-up and treatment outcome durability is almost universally lacking in the literature and only 64.8% of patients have reported outcome data. Combining preoperative diagnoses further limits the conclusions able to be drawn as life expectancy for some conditions, i.e. those with oncological facial pain, may be shorter than the time it may take to achieve full benefit from the procedure.

## Conclusion

This review summarizes 80 years of literature regarding interventions targeting the NC for chronic refractory craniofacial pain. Recent advancements in intervention techniques are highlighted, but also shortcomings in the literature impede widespread adoption. Only 64.8% of included subjects have outcomes reported and due to the inclusion of studies prior to the standardization of pain reporting metrics, there are inconsistencies in the way these outcomes are reported. New and less invasive techniques continue to emerge, however prospective studies remain absent in the literature. Additional studies addressing efficacy comparisons between intervention type or preoperative diagnosis would help solidify these techniques in the armamentarium of craniofacial pain treatments.

## Data Availability

All data produced in the present study are available upon reasonable request to the authors.

