## Supplemental Material 1 for "Surgical interventions targeting the nucleus caudalis for craniofacial pain: a systematic and historical review"

EMBASE (Ovid database; Search date: May 31, 2021)

1. Caudalis.mp.
2. trigeminal nucleus.mp.
3. trigeminal tractotom*.mp.
4. trigeminal Nucleotractotom*.mp.
5. trigeminal nucleotom*.mp.
6. Dorsal root entry zone.mp.
7. Drez.mp.
8. Ablat*.mp.
9. Lesion.mp.
10. Stimulat*.mp.
11. 6 or 7 or 8 or 9 or 10
12. 1 or 2
13. 11 and 12
14. 3 or 4 or 5 or 13
15. Limit to Humans

Results= 519

Medline (Ovid database; Search date: May 31, 2021)

1. Caudalis.mp.
2. trigeminal nucleus.mp.
3. trigeminal tractotom*.mp.
4. trigeminal Nucleotractotom*.mp.
5. trigeminal nucleotom*.mp.
6. Dorsal root entry zone.mp.
7. Drez.mp.
8. Ablat*.mp.
9. Lesion.mp.
10. Stimulat*.mp.
11. 6 or 7 or 8 or 9 or 10
12. 1 or 2
13. 11 and 12
14. 3 or 4 or 5 or 13
15. Limit to Humans

Results= 249

Cochrane CENTRAL database (Search date: May 31, 2021)

1. Caudalis.mp.
2. “trigeminal nucleus”
3. trigeminal NEXT tractotom*
4. trigeminal NEXT Nucleotractotom*
5. trigeminal NEXT nucleotom*
6. “Dorsal root entry zone”
7. Drez
8. Ablat*
9. Lesion
10. Stimulat*
11. 6 or 7 or 8 or 9 or 10
12. 1 or 2
13. 11 and 12
14. 3 or 4 or 5 or 13

Results= 14
