## Supplemental Material 2 for "Surgical interventions targeting the nucleus caudalis for craniofacial pain: a systematic and historical review"

**Supplementary Material 2:** Summary of articles reporting interventions targeting the nucleus caudalis for facial pain with fewer than 5 included patients.

| Author (year) | Patients, # | Diagnoses (n) | Intervention Technique | Fraction of patients improved |
| --- | --- | --- | --- | --- |
| Todd (1969)^1^ | 3 | TN (3) | Percutaneous nucleotomy/tractotomy | 100% |
| Hitchcock (1972)^2^ | 3 | Postherpetic (3) | Open DREZ nucleotomy/tractotomy | 100% |
| Siqueira (1985)^3^ | 2 | Postherpetic (1)  Anesthesia Dolorosa (1) | Open DREZ nucleotomy/tractotomy | 100% |
| Hitchcock (1987)^4^ | 4 | Not specified (4) | Open DREZ nucleotomy/tractotomy | 75% |
| Ishijima (1988)^5^ | 4 | Postherpetic (4) | Open DREZ nucleotomy/tractotomy | 100% |
| Spiegelmann (1991)^6^ | 2 | Postherpetic (1)  Anesthesia Dolorosa (1) | Open DREZ nucleotomy/tractotomy | 100% |
| Sampson (1992)^7^ | 2 | Post Stroke (2) | Open DREZ nucleotomy/tractotomy | 100% |
| Chang (2002)^8^ | 2 | TN (2) | Gamma Knife Radiosurgery | 100% |
| Teixeira (2007)^9^ | 2 | Anesthesia Dolorosa (2) | Percutaneous nucleotomy/tractotomy | 100% |
| Samreen (2009)^10^ | 1 | Postherpetic (1) | Open DREZ nucleotomy/tractotomy | 100% |
| Teixeira (2012)^11^ | 1 | Postherpetic (1) | Endoscopic DREZ nucleotomy/tractotomy | 100% |
| Sandwell (2013)^12^ | 1 | Anesthesia Dolorosa (1) | Open DREZ nucleotomy/tractotomy | 100% |
| Thompson (2013)^13^ | 2 | Postherpetic (1)  Anesthesia Dolorosa (1) | Percutaneous nucleotomy/tractotomy | 100% |
| Yearwood (2016)^14^ | 1 | Headache (1) | Cervical SCS | 100% |
| Kanpolat (2017)^15^ | 1 | Not specified (1) | Open DREZ nucleotomy/tractotomy | 100% |
| Richter (2017)^16^ | 1 | TN (1) | Cervical SCS | 100% |
| Jones (2020)^17^ | 1 | Traumatic (1) | Cervical SCS | 100% |

Abbreviations: DREZ, dorsal root entry zone; SCS, spinal cord stimulation; TN, Trigeminal Neuralgia/trigeminal neuropathic pain.
