## Supplemental Material 3 for "Surgical interventions targeting the nucleus caudalis for craniofacial pain: a systematic and historical review"

**Supplemental Material 2**

Table 1: Analysis of outcomes between intervention techniques.

|  | Composite Outcome Score  Proportion [95% CI] | | | | Statistics | | |
| --- | --- | --- | --- | --- | --- | --- | --- |
|  | 1 | 2 | 3 | 4 | | χ2 | p-value |
| Focal Lesion (nucleotomy/tractotomy) | 0.55 [0.48, 0.63] | 0.22 [0.17, 0.30] | 0.12 [0.07, 0.18] | 0.10 [0.06, 0.16] | | 57.5322 | <0.001 |
| DREZ | 0.31 [0.24, 0.39] | 0.50 [0.42, 0.59] | 0.06 [0.03, 0.11] | 0.13 [0.08, 0.19] | |  |  |
| Percutaneous nucleotomy/tractotomy | 0.48 [0.41, 0.54] | 0.42 [0.36, 0.49] | 0.04 [0.02, 0.08] | 0.06 [0.04, 0.10] | |  |  |
| Neuromodulation (SCS) | 0.03 [0.00, 0.32] | 0.88 [0.63, 0.97] | 0.03 [0.00, 0.32] | 0.12 [0.03, 0.37] | |  |  |
