## Supplemental Material 4 for "Surgical interventions targeting the nucleus caudalis for craniofacial pain: a systematic and historical review"

**Supplemental Material 3**

**Table 1:** Correlation estimates for the fraction of patients demonstrating postoperative improvement with respect to publication date. Abbreviations: DREZ, dorsal root entry zone.

|  | Covariance | Correlation Coefficient | P-value |
| --- | --- | --- | --- |
| All interventions | -0.01 | -0.003 | 0.99 |
| Open Focal Lesioning | 1.05 | 0.45 | 0.19 |
| Open DREZ | -0.13 | -0.09 | 0.80 |
| Percutaneous | 1.12 | 0.44 | 0.33 |

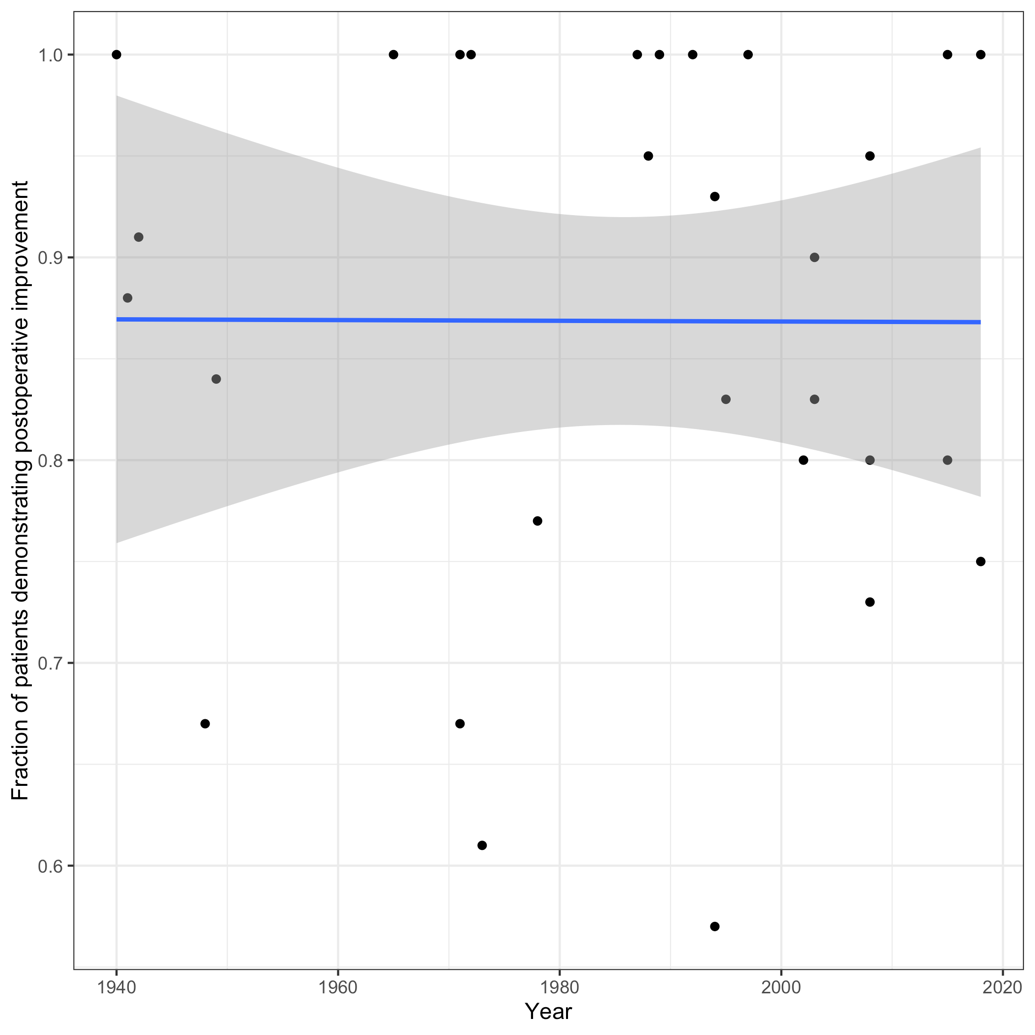

**Figure 1: Correlation between fraction of patients demonstrating postoperative improvement and publication year for all intervention types.** There is no correlation between these two factors as demonstrated by the plotted linear model (blue) and standard error (grey).

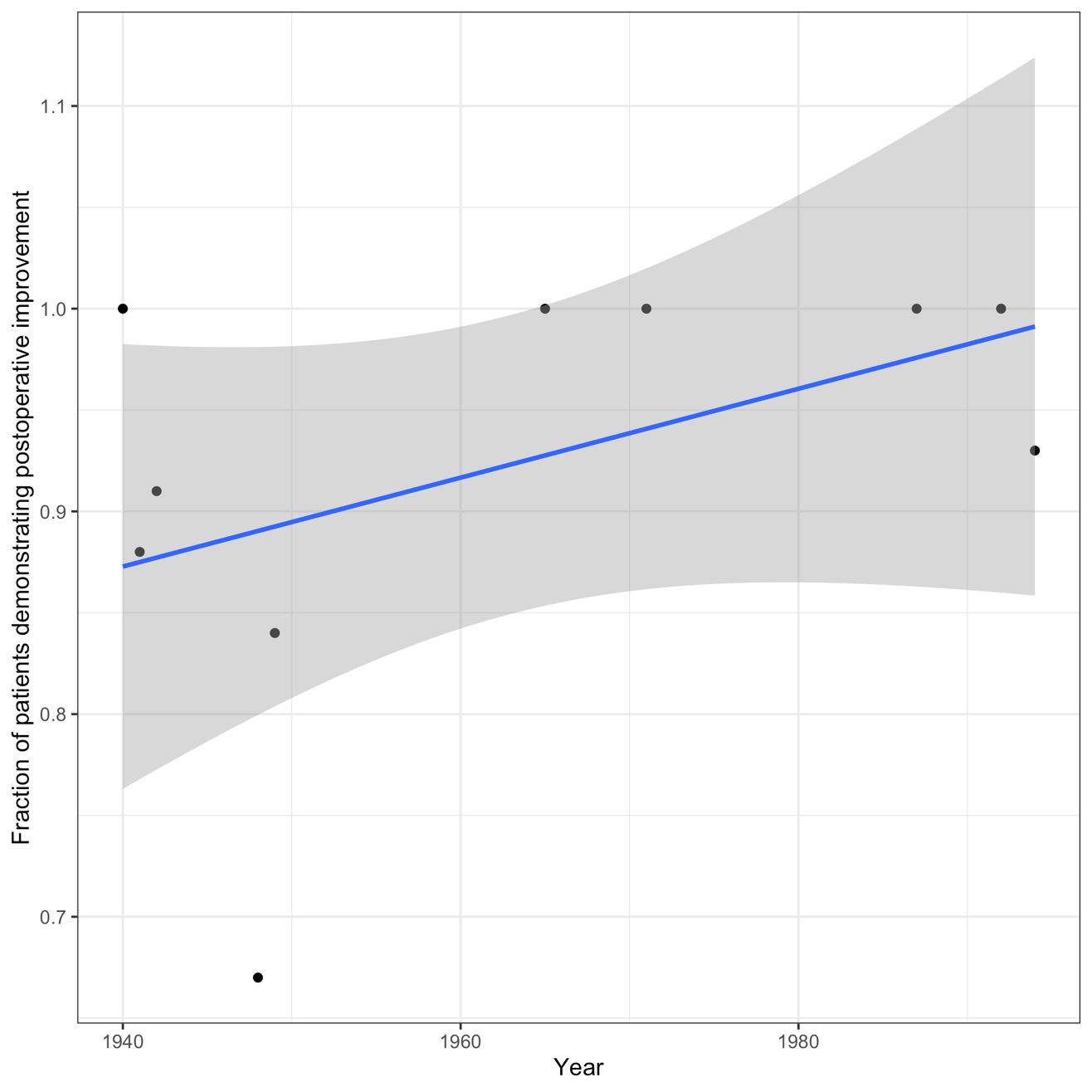

**Figure 2: Correlation between fraction of patients demonstrating postoperative improvement and publication year for open focal lesioning techniques (nucleotomy/tractotomy).** There is a slight linear correlation between these two factors as demonstrated by the plotted linear model (blue) and standard error (grey).

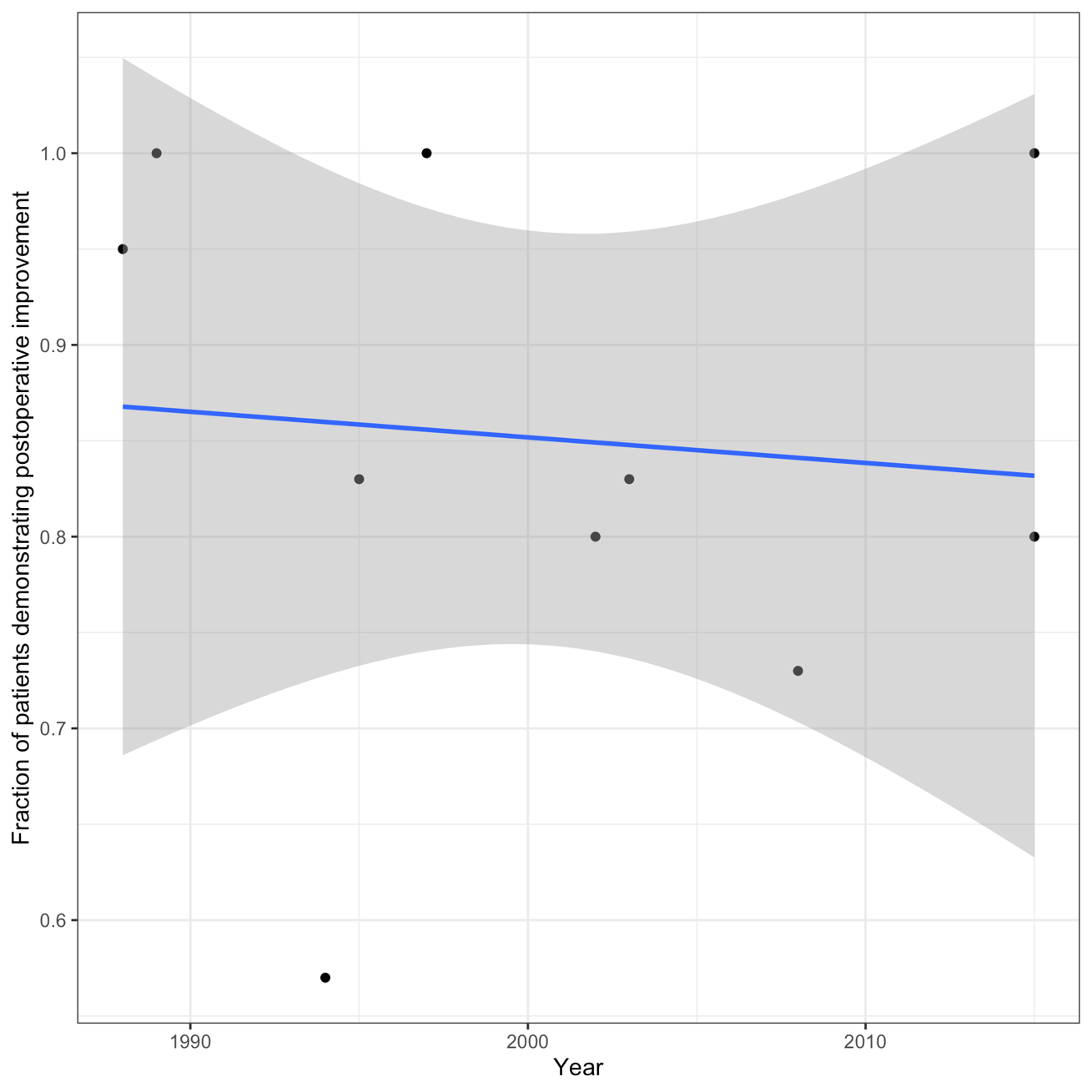

**Figure 3: Correlation between fraction of patients demonstrating postoperative improvement and publication year for open DREZ (multipoint lesioning technique).** There is a slight negative linear correlation between these two factors as demonstrated by the plotted linear model (blue) and standard error (grey). Abbreviations: DREZ, dorsal root entry zone.

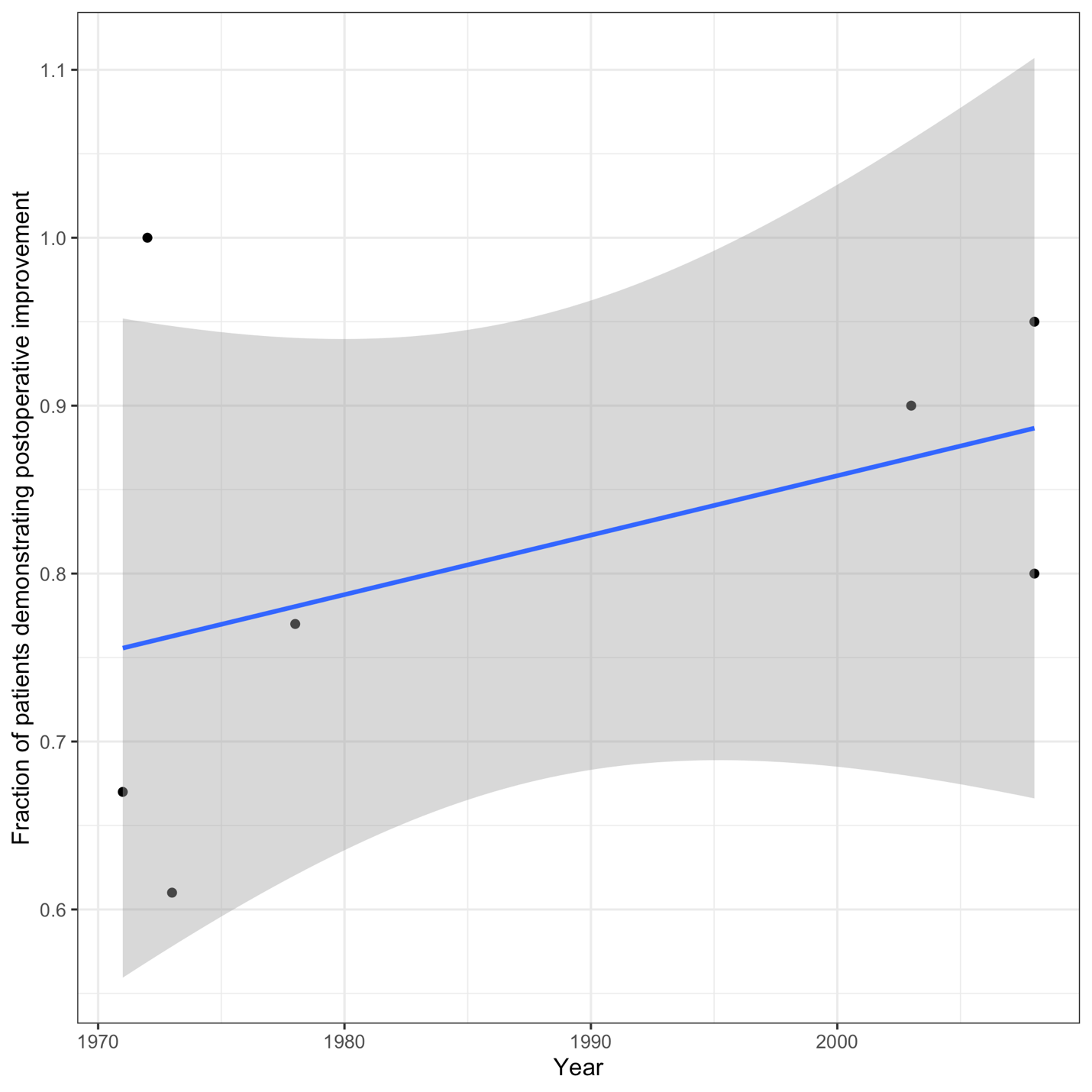

**Figure 4: Correlation between fraction of patients demonstrating postoperative improvement and publication year for percutaneous nucleotomy/tractotomy.** There is a slight linear correlation between these two factors as demonstrated by the plotted linear model (blue) and standard error (grey).
