## Supplemental Material 5 for "Surgical interventions targeting the nucleus caudalis for craniofacial pain: a systematic and historical review"

**Supplemental Material 4**

**Table 1:** Analysis of complications between intervention techniques. Numbers are proportion followed by [95% CI].

|  |  | Statistics | |  | Statistics | |
| --- | --- | --- | --- | --- | --- | --- |
|  | Facial pain/numbness/weakness | χ2 | p-value | Corneal Anesthesia | χ2 | p-value |
| Focal Lesion (Nucleotomy/tractotomy) | 0.09 [0.06, 0.13] | 46.13 | <0.001 | 0.01 [0.00, 0.02] | 5.44 | 0.142 |
| DREZ | 0.00 [0.00, 0.04] |  |  | 0.02 [0.01, 0.05] |  |  |
| Percutaneous nucleotomy/tractotomy | 0.00 [0.00, 0.03] |  |  | 0.00 [0.00, 0.03] |  |  |
| Neuromodulation (SCS) | 0.03 [0.00, 0.32] |  |  | 0.03 [0.00, 0.32] |  |  |
|  |  | Statistics | |  | Statistics | |
|  | Oropharyngeal Dysfunction | χ2 | p-value | Ataxia | χ2 | p-value |
| Focal Lesion (Nucleotomy/tractotomy) | 0.03 [0.02, 0.05] | 11.52 | 0.009 | 0.06 [0.04, 0.09] | 41.36 | <0.001 |
| DREZ | 0.00 [0.00, 0.04] |  |  | 0.14 [0.10, 0.20] |  |  |
| Percutaneous nucleotomy/tractotomy | 0.00 [0.00, 0.02] |  |  | 0.00 [0.00, 0.02] |  |  |
| Neuromodulation (SCS) | 0.02 [0.00, 0.32] |  |  | 0.03 [0.00, 0.32] |  |  |
|  |  | Statistics | |  | Statistics | |
|  | Limb Dysfunction | χ2 | p-value | Transient Dysfunction | χ2 | p-value |
| Focal Lesion (Nucleotomy/tractotomy) | 0.01 [0.00, 0.03] | 12.08 | 0.007 | 0.06 [0.04, 0.09] | 17.42 | <0.001 |
| DREZ | 0.00 [0.00, 0.04] |  |  | 0.15 [0.11, 0.21] |  |  |
| Percutaneous nucleotomy/tractotomy | 0.04 [0.02, 0.07] |  |  | 0.06 [0.04, 0.10] |  |  |
| Neuromodulation (SCS) | 0.03 [0.00, 0.32] |  |  | 0.03 [0.00, 0.32] |  |  |
|  |  | Statistics | |  | Statistics | |
|  | Reoperation | χ2 | p-value | General Medical | χ2 | p-value |
| Focal Lesion (Nucleotomy/tractotomy) | 0.04 [0.02, 0.06] | 75.27 | <0.001 | 0.01 [0.00, 0.03] | 21.86 | <0.001 |
| DREZ | 0.00 [0.00, 0.04] |  |  | 0.05 [0.03, 0.10] |  |  |
| Percutaneous nucleotomy/tractotomy | 0.03 [0.02, 0.06] |  |  | 0.00 [0.00, 0.03] |  |  |
| Neuromodulation (SCS) | 0.41 [0.21, 0.65] |  |  | 0.03 [0.00, 0.32] |  |  |
|  |  | Statistics | |  | Statistics | |
|  | Mortality | χ2 | p-value | Total | χ2 | p-value |
| Focal Lesion (Nucleotomy/tractotomy) | 0.06 [0.04, 0.09] | 27.71 | <0.001 | 0.38 [0.33, 0.43] | 46.97 | <0.001 |
| DREZ | 0.01 [0.00, 0.04] |  |  | 0.40 [0.33, 0.47] |  |  |
| Percutaneous nucleotomy/tractotomy | 0.00 [0.00, 0.03] |  |  | 0.16 [0.12, 0.20] |  |  |
| Neuromodulation (SCS) | 0.02 [0.00, 0.32] |  |  | 0.41 [0.21, 0.65] |  |  |
